# Mitochondrial DNA copy number and trimethylamine levels in the blood: new insights on cardiovascular disease biomarkers

**DOI:** 10.1101/2020.12.29.20248667

**Authors:** Laura Bordoni, Irene Petracci, Iwona Pelikant-Malecka, Adriana Radulska, Marco Piangerelli, Joanna J. Samulak, Lukasz Lewicki, Leszek Kalinowski, Rosita Gabbianelli, Robert A. Olek

## Abstract

Among cardiovascular disease (CVD) biomarkers, the mitochondrial DNA copy number (mtDNAcn) is a promising candidate. A growing attention has been also dedicated to trimethylamine-N-oxide (TMAO), an oxidative derivative of the gut metabolite trimethylamine (TMA). With the aim to identify biomarkers predictive of CVD, we investigated TMA, TMAO and mtDNAcn in a population of 389 coronary artery disease (CAD) patients and 151 healthy controls, in association with established risk factors for CVD (gender, age, hypertension, smoking, diabetes, glomerular filtration rate (GFR)). MtDNAcn was significantly lower in CAD patients and in hypertensive subjects; it correlates with GFR and TMA, but not with TMAO. A biomarker including mtDNAcn, gender, and hypertension (but neither TMA nor TMAO) emerged as a good predictor of CAD. Our findings support the usage of mtDNAcn as a plastic biomarker to monitor the exposure to risk factors and the efficacy of preventive interventions for a personalized CAD risk reduction.

**Highlights:**

- mtDNAcn measured in whole blood is associated to the cardiovascular health status in humans;
- mtDNAcn is reduced in CAD and hypertension, and inversely correlates with GFR;
- mtDNA, gender and hypertension together represent a good predictive biomarker for CAD;
- TMA metabolism is different in healthy and CAD subjects;
- TMA and TMAO are not good predictors of CAD.

## INTRODUCTION

Cardiovascular disease (CVD) is a major cause of death and disability worldwide (Roth et al., 2017). The primary prevention of CVD relies on the identification of high-risk individuals before the manifestation of the event. Thus, the need for new methods for accurate risk stratification is drawing attention, and several biomarkers have been proposed to predict cardiovascular events (Wang et al., 2017). Biomarkers play a critical role in the definition, prognosis, and decision-making in cardiovascular disease management. Nevertheless, despite increasing efforts, the possibility to identify high-risk individuals is still limited. Among the novel candidates, trimethylamine N-oxide (TMAO), a metabolite of the gut microbe-derived trimethylamine (TMA), has attracted a growing attention as a potential promoter of atherosclerosis in humans (Kanitsoraphan et al., 2018; Koeth et al., 2013; Senthong et al., 2016; Tang et al., 2013; Wang et al., 2011). Several hypotheses have linked the TMAO to the development of atherosclerosis and CVD risk via promotion of platelet hyper-reactivity (Zhu et al., 2016), proinflammatory changes in the artery wall (Seldin et al., 2016), enhanced macrophage cholesterol accumulation and foam cell formation (Wang et al., 2011), increased levels of proinflammatory monocytes (Haghikia et al., 2018). Nevertheless, contrasting evidence on the association of TMAO and CVD emerged (Bordoni et al., 2020c; Collins et al., 2016; Jia et al., 2020; Landfald et al., 2017; Olek et al., 2019; Samulak et al., 2019), and a clear mechanistic explanation of this association is still missing.

Another interesting peripheral biomarker of CVD that has been recently proposed is the evaluation of mitochondrial DNA copy number (mtDNAcn) (Ashar et al., 2017). Human mitochondrial DNA (mtDNA) is a small, circular, and multi-copy genome, located in the inner matrix of mitochondria. It incorporates 37 mitochondrial genes (13 coding for essential components of the mitochondrial electron transport chain and of the ATP synthase complex, 22 for mitochondrial transfer RNAs and 2 for ribosomal RNAs). Mitochondrial DNA content reflects the energy demand of a cell (Melser et al., 2015) and is disturbed by imbalanced energy metabolism and reactive oxygen species overproduction. Thus, mtDNAcn changes have been proposed as an early biomarker of damage and mitochondrial dysfunction (Clay Montier et al., 2009; Malik and Czajka, 2013). Remarkably, since mtDNAcn changes have been associated with both intrinsic and extrinsic factors (Bordoni and Gabbianelli, 2020; Torres, 2018), mtDNAcn has been proposed as a potential biomarker for complex diseases, which are linked to both genetics and environmental exposures (Castellani et al., 2020).

Circulating mtDNAcn has been investigated in cardiovascular diseases (Yue et al., 2018), mainly analyzing peripheral blood cells. Interestingly, it has been recently demonstrated that blood also contains circulating cell-free respiratory competent mitochondria (Al Amir Dache et al., 2020), suggesting that the measure of mtDNAcn in the whole blood might better represent the health status. Moreover, despite the involvement of TMAO in mitochondrial metabolism has been presented (Chen et al., 2017; Makrecka-Kuka et al., 2017; Schneider et al., 2018; Vallance et al., 2018), none of the previous investigations examined the association between blood mtDNAcn and TMA or TMAO levels in humans.

This study aimed to investigate, in a population of 540 subjects of coronary artery disease (CAD) patients and controls, 1) if the mtDNA copy number measured in the whole blood is a marker of CAD; 2) if any association between mtDNAcn and TMAO or TMA levels exists; 3) if mtDNAcn is associated with other risk factors for CVD that are linked to metabolic alterations (i.e. hypertension (Fuchs and Whelton, 2020), diabetes (Leon and Maddox, 2015), glomerular filtration rate (GFR)) or environmental exposures (i.e. smoking (Barbara and David, 2014), BMI (Khan et al., 2018)); 4) provide further insights on the effects of TMA and TMAO levels changes in CVD. The final aim is to identify relevant factors that can be used for CVD prognosis through adequate prediction models.

## RESULTS

### Descriptive statistics

540 recruited subjects (65.7% male, 34.3% female) were analyzed in this study. Mean age of the population was 65 (±10) years old. Among the subjects, 46.7% (n=252) were smokers, 66.5% (n=359) were diagnosed with hypertension, and 25.4% (n=137) were diagnosed with diabetes. Among the recruited subjects, 151 were controls (28%), 389 were CAD patients (72%). Descriptive statistics for BMI, TMA, TMAO, TMAO/TMA ratio and GFR in the total population are shown in table 1.

**Table 1.** Descriptive statistics on the whole population (n=540) for the analyzed variables. SD: standard deviation.

|  | n | Min | Max | Mean | SD |
| --- | --- | --- | --- | --- | --- |
| **BMI (kg/m<sup>2</sup>)** | 540 | 18.00 | 45.00 | 28.50 | 4.40 |
| **TMAO (μM)** | 540 | 0.01 | 37.50 | 5.37 | 4.50 |
| **TMA (μM)** | 540 | 0.34 | 1.15 | 0.60 | 0.11 |
| **TMAO/TMA** | 540 | 0.01 | 64.87 | 8.86 | 7.35 |
| **GFR (ml/min/1.73m<sup>2</sup>)** | 540 | 9.71 | 289.96 | 88.17 | 33.76 |

### Risk factors for CAD distribution in the analyzed population

As expected, smoking (Pearson’s chi-square=4.86; p=0.028), hypertension (Pearson’s chi-square=60.79; p=0.0001), and diabetes (Pearson’s chi-square=14.55; p=0.0001) are confirmed as risk factors for CAD development (figure 1 supplementary materials). An increased BMI was also measured in CAD patients with respect to controls (controls: 27.8±4.1; CAD: 28.8±4.5; F=5.18; p=0.023). A detailed comparison of TMA, TMAO, BMI and GFR in the CAD population vs controls in this population has been previously published (Bordoni et al., 2020a).

**Figure 1.**
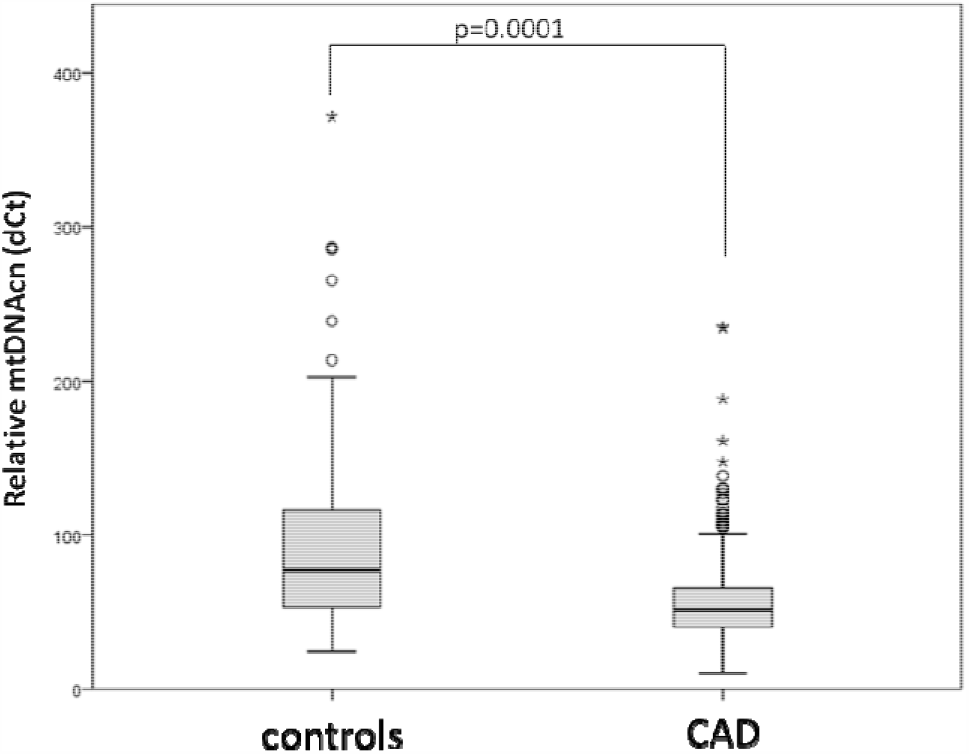
mtDNAcn in controls and CAD group.

### Association between mtDNAcn, CAD and CVD risk factors

In the studied population, blood mtDNAcn decreased with age (Spearman’s rho=-0.101; p= 0.019) (figure 2A supplementary materials). A significantly lower level of mtDNAcn was measured in CAD group compared to controls (median values in controls vs CAD: 76.9 vs 51.5; F=107.65; p=0.0001) (figure 1). Analysis of covariates (age, gender, TMA, TMAO, GFR) revealed that this association is modulated by TMA (F=14.94; p=0.0001) and GFR (F=4.97; p=0.026).

**Figure 2.**
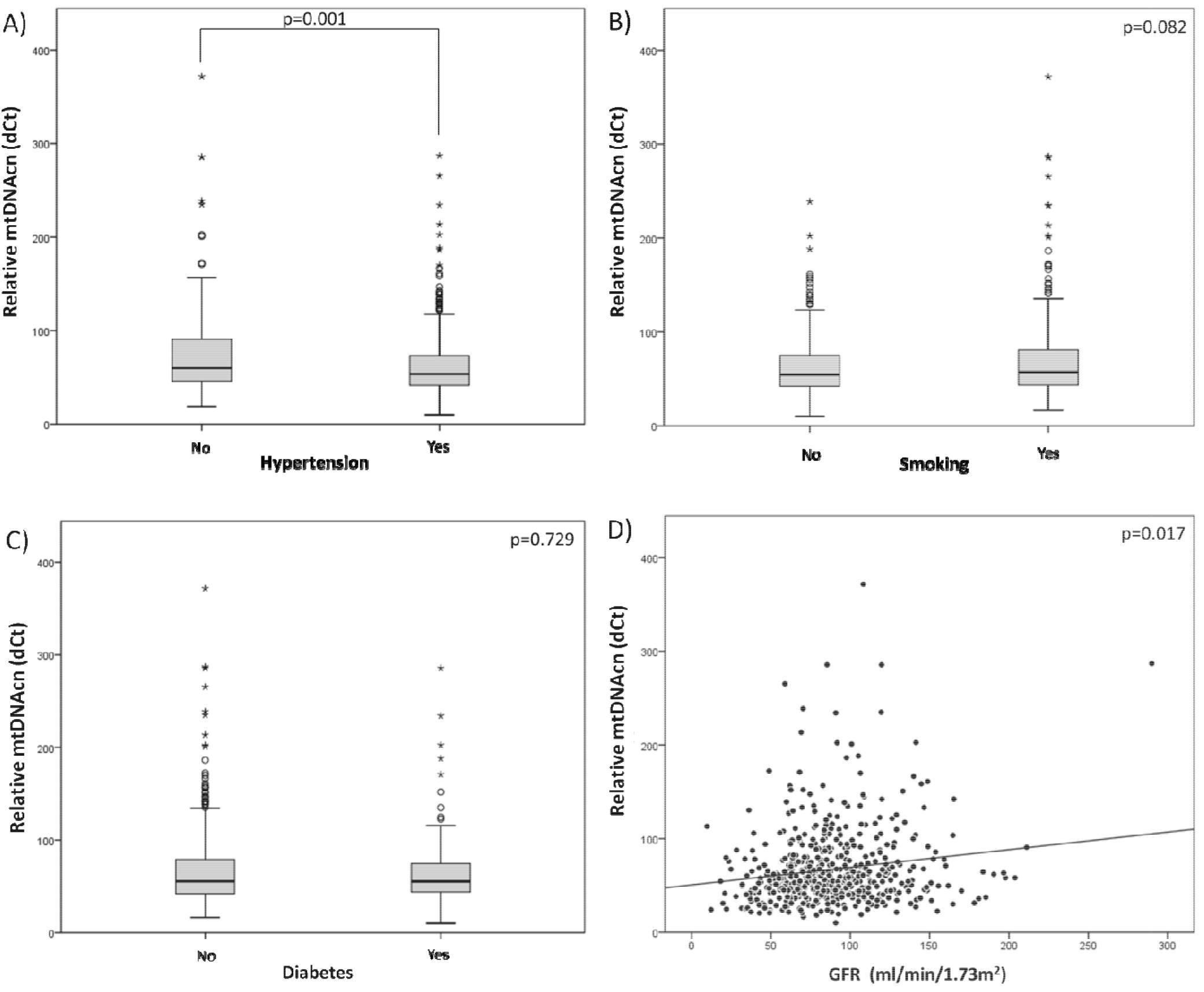
MtDNAcn and CAD risk factors exposures in the analyzed population: hypertension (A), smoking (B), diabetes (C), GFR (D)

Moreover, mtDNAcn was significantly lower in individuals with diagnosed hypertension (median values, control=59.8; CAD = 53.6; F=10.03; p=0.001) (Fig. 2A). This association is modulated by TMA (F=22.53; p=0.0001) and GFR (F=9.75; p=0.002). MtDNAcn did not differ in individuals diagnosed with diabetes (F=0.12; p=0.729) (Fig. 2B), nor in smokers with respect to controls (F=3.04; p=0.082) (figure 2C). MtDNAcn was directly correlated with GFR (Spearman’s rho=0.103; p=0.017), suggesting that higher mtDNAcn can be associated with a better glomerular filtration capacity (figure 2D). However, this correlation is only nominal (p=0.068) after Bonferroni’s correction. No significant association was measured between mtDNAcn and BMI in the whole population (Spearman’s rho=0.044; p= 0.304).

### MtDNA correlates with TMA but not TMAO

A direct correlation between mtDNAcn and TMA levels was measured (Spearman’s rho=0.166; p=0.0001) (figure 3A). This correlation is still significant (p=0.0004) after Bonferroni’s correction. No significant association between mtDNAcn and TMAO (Spearman’s rho=0.020; p=0.643) (figure 3B) or TMAO/TMA (Spearman’s rho=-0.033; p=0.441) (figure 3C) has been detected in the total population.

**Figure 3.**
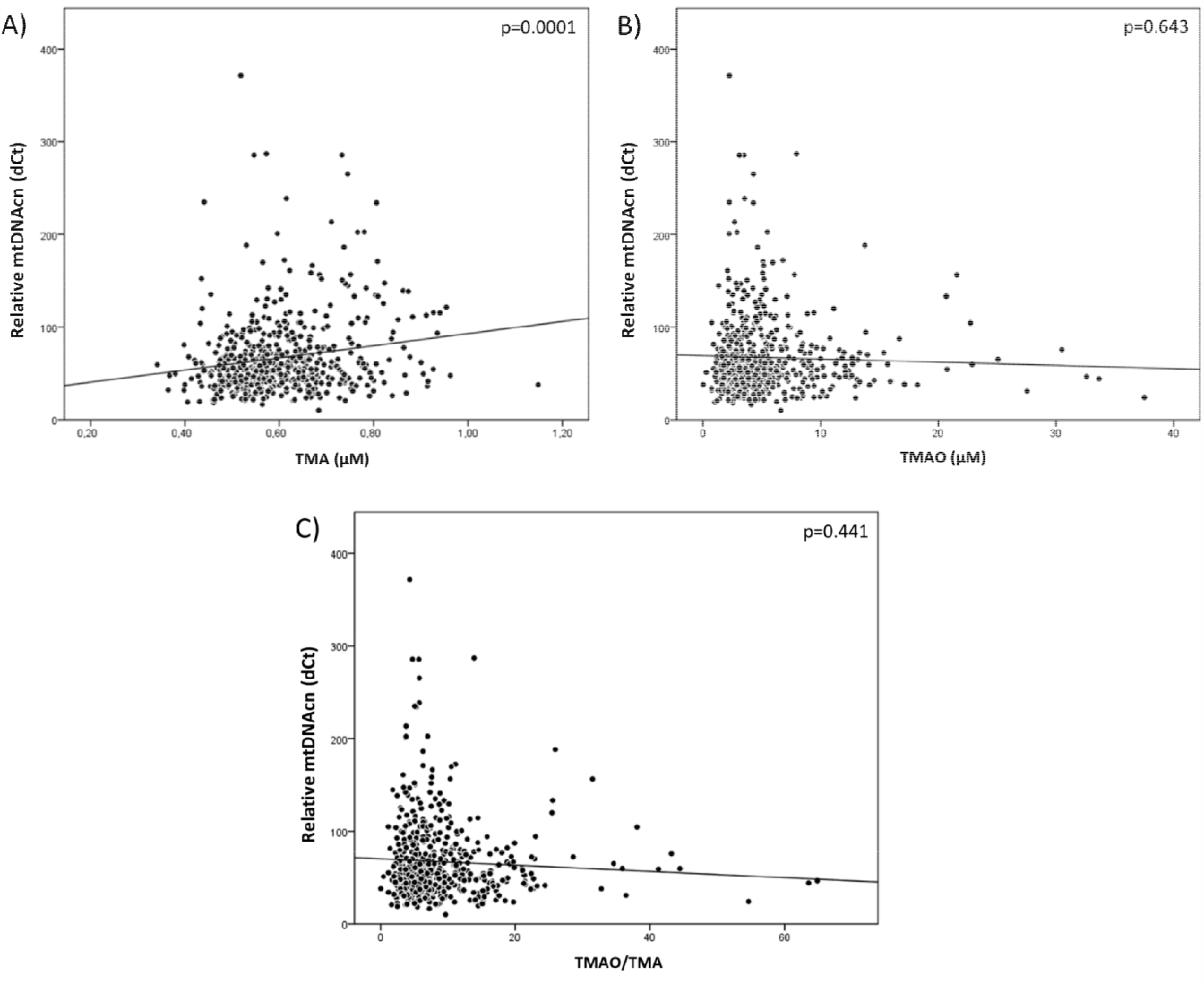
Correlations between mtDNAcn and TMA (A), TMAO (B) or TMAO/TMA (C) in the analyzed population

### Correlations between TMA, TMAO, GFR and mtDNAcn in controls and CAD patients

To clarify the role of TMA in this complex picture, we performed an exploratory analysis to test the correlations between TMA and TMAO in the control and CAD groups separately. We observed that, while high TMA levels are associated with high TMAO levels in CAD patients (Spearman’s rho=0.338; p=0.0001), high TMA levels are not accompanied by increased TMAO in controls (Spearman’s rho= 0.065; p=0.429) (figure 4A). This corroborates the hypothesis that TMA is not *per se* linked to high TMAO levels. Coherently, it is possible to observe that higher TMA levels are positively correlated to GFR in the controls (Spearman’s rho= 0.257; p=0.001), while TMA is negatively associated to GFR levels in the CAD group (Spearman’s rho=-0.260; p=0.0001) (figure 4B). Moreover, increased TMA levels correlated to higher mtDNAcn only in the controls (Spearman’s rho=0.280, p=0.0001), but not in the CAD group (Spearman’s rho = 0.076, p=0.133) (figure 4C). This suggests a different metabolism of TMA and TMAO in the presence of cardiovascular disease, with respect to healthy conditions.

**Figure 4.**
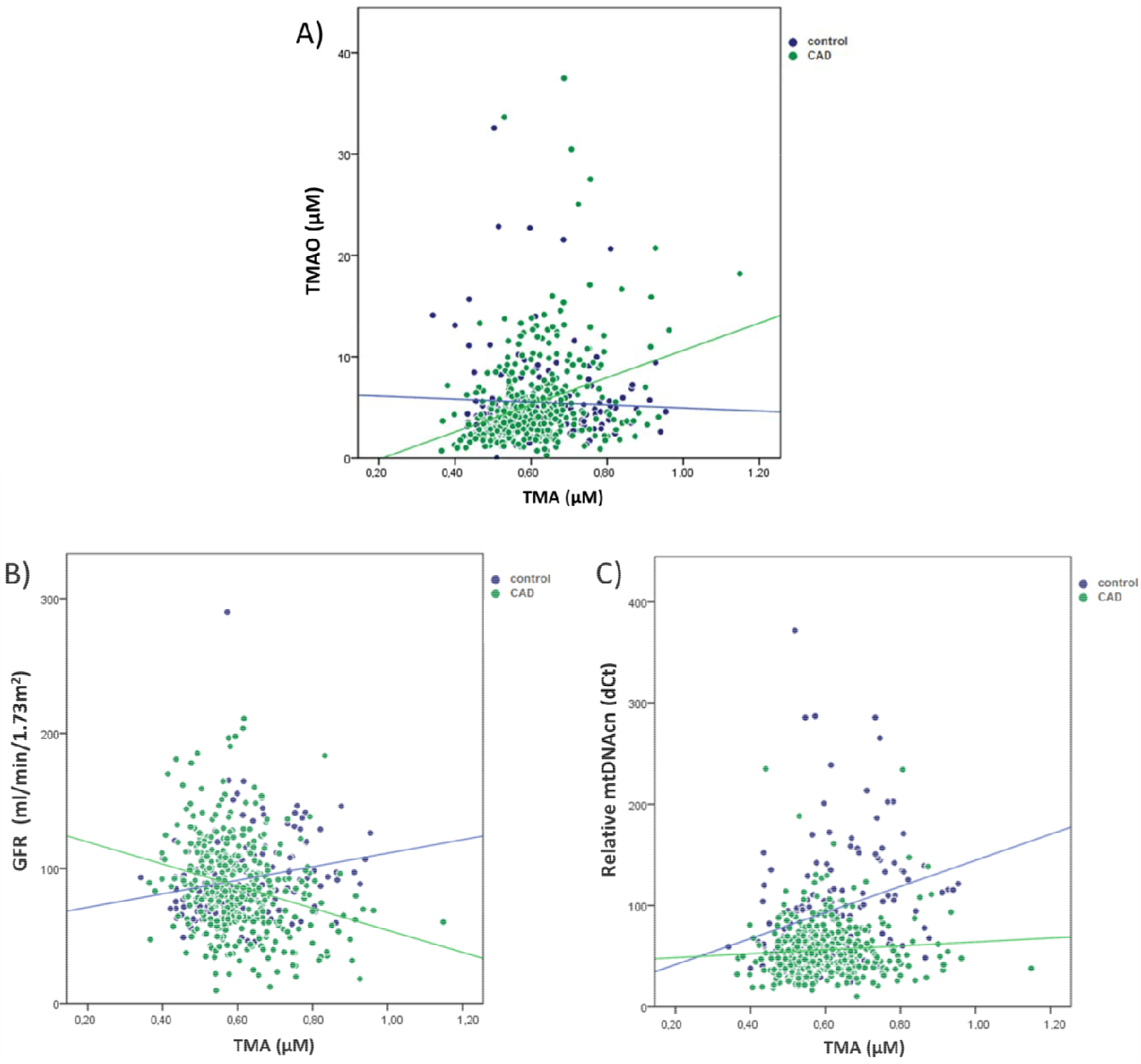
Correlations between TMA and TMAO (A), GFR (B), and mtDNAcn (C) in the population divided for groups (controls vs CAD).

By observing the TMAO/TMA ratio in the population divided by the cardiovascular health status in respect to mtDNAcn (Figure 5A) and GFR (Figure 5B), we found that the TMAO/TMA ratio is inversely correlated to mtDNAcn in the control group (Spearman’s rho = −0.161; p= 0.049), but not in the CAD group (Spearman’s rho = −0.019; p = 0.712). Moreover, the TMAO/TMA ratio shows a significant inverse correlation with GFR in the CAD group (Spearman’s rho = −0.307; p = 0.0001), which is weaker in the controls (Spearman’s rho = −0.176; p = 0.030).

**Figure 5.**
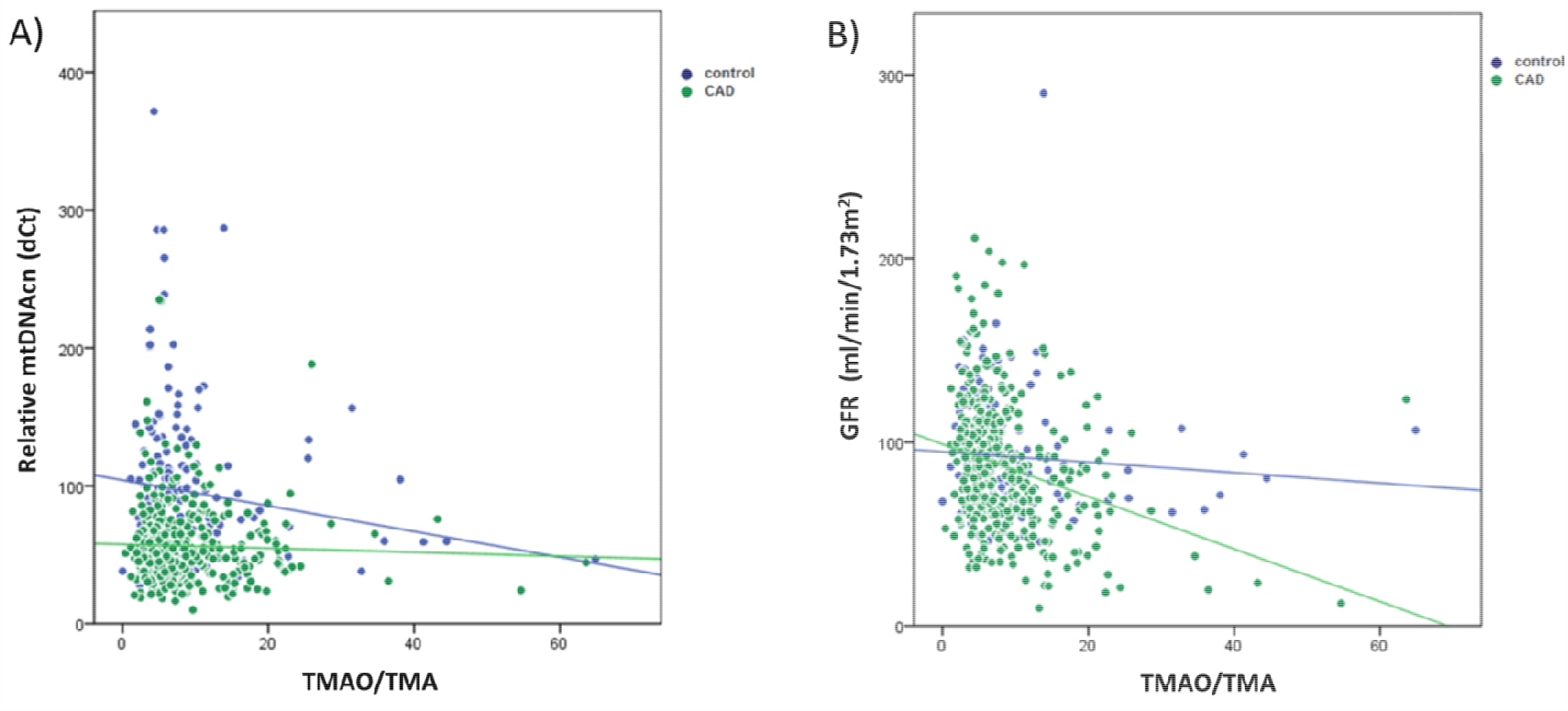
Correlations between TMAO/TMA ratio and mtDNAcn (A) and GFR (B) in the population divided for groups (controls vs CAD).

### CAD prediction models

The stepwise logistic regression analysis identified the following variables as significant predictors of CAD: mtDNAcn (Exp(B)=0.972; 95%CI=[0.965-0.979]; p=0.0001;); hypertension (Exp(B)=4.131; 95%CI=[2.582-6.609]p=0.0001;); sex (Exp(B)=0.574; 95%CI=[0.355-0.929]; p=0.024); smoking (Exp(B)=1.754; 95%CI=[1.083-2.841]; p=0.022); diabetes (Exp(B)=1.861; 95%CI=[1.011-3.425]; p=0.045,). TMAO and TMA are not established as significant predictors of CAD in this model. On the contrary, mtDNAcn is the first element to enter in the stepwise analysis, thus affecting the goodness of the prediction and the information carried by the other biomarkers.

The precision-recall (PR) analysis was applied to evaluate the performance of the prediction of 2 models (3-step and 5-step models) identified by the stepwise logistic regression. PR shows an area under the curve (AUC) = 0.894 for the 3-step model, where mtDNAcn, hypertension and sex were the predictors. For the 5-step model (where mtDNAcn, hypertension, sex, smoking, diabetes were the predictor), the resulting AUC was 0.901. Figure 6 shows the precision-recall curve for both models, concluding that their performances are good and similar.

**Figure 6.**
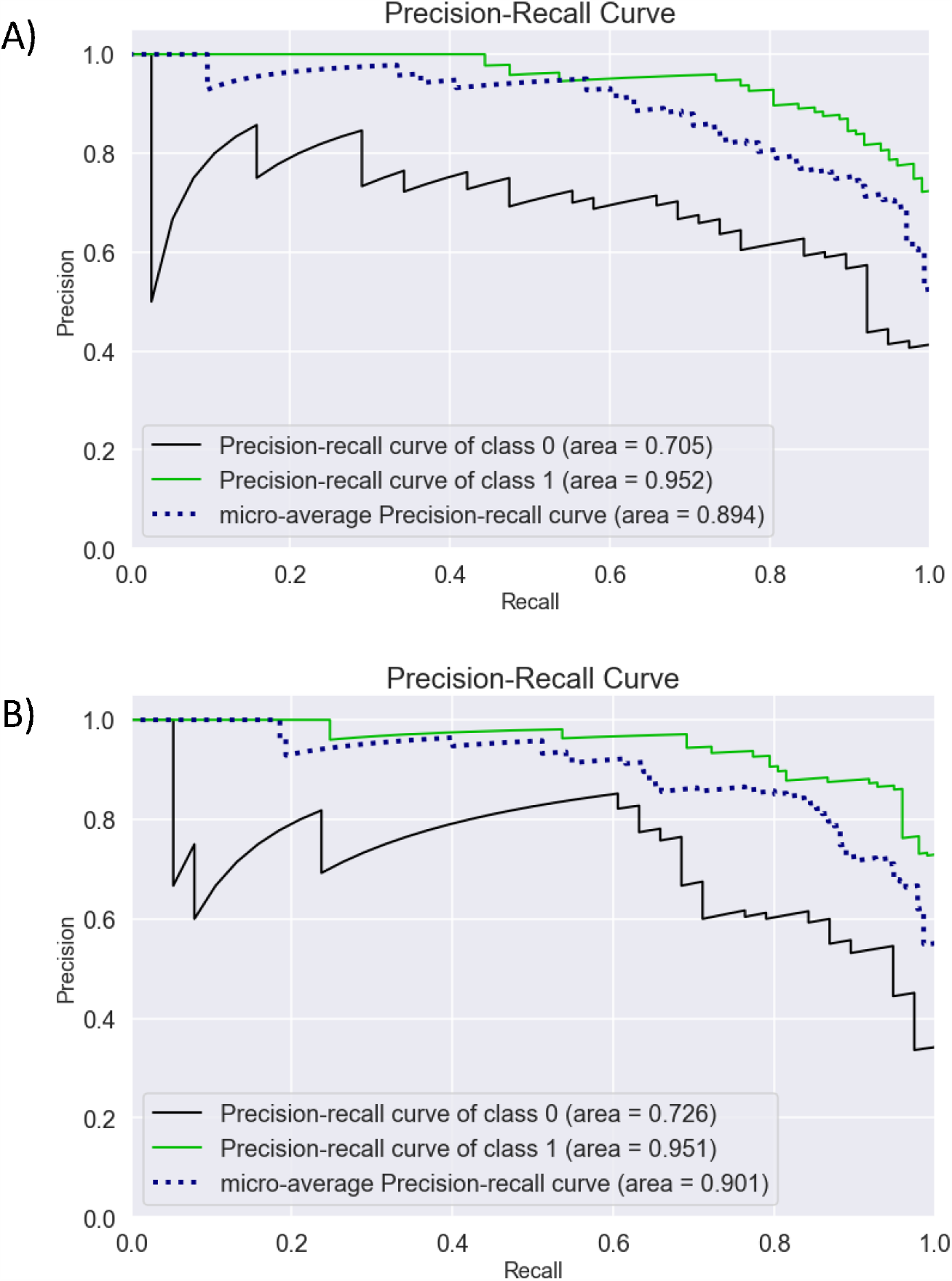
Precision-recall analysis for CAD prediction with 3 features (A, 3-step model: including mtDNAcn, hypertension, sex) or 5 features (B, 5-step model: including mtDNAcn, hypertension, sex, smoking, diabetes).

## DISCUSSION

Alterations in TMAO metabolism (Velasquez et al., 2016) and mitochondrial dynamics (Silva et al., 2016) have been previously independently identified as risk factors for cardiovascular disease development. We have found that mtDNA copy number, measured in the whole blood, is lower in CAD patients than in controls. This is coherent with previous investigations reviewed by Yue et al. (Yue et al., 2018); although, the reported studies had determined mtDNAcn in buffy coat/circulating leukocytes. Recently, it has been demonstrated that blood contains circulating cell-free respiratory competent mitochondria (Al Amir Dache et al., 2020), and a reduction of mtDNAcn was also detected in the whole blood of peripheral arterial disease male patients compared to controls (Koller et al., 2020). Therefore, our findings support the interest towards mtDNAcn measurement in the whole blood as an easy-accessible biomarker for arterial disease risk stratification.

In our population, mtDNAcn decreased with age, according to previous evidence examining mtDNAcn in blood cells (Mengel-From et al., 2014). In addition, we found a correlation between mtDNAcn, hypertension and GFR, which are established risk factors for CVD (Flint et al., 2019; Manjunath et al., 2003; Yidan et al., 2018). In particular, mtDNAcn was decreased in hypertensive subjects. Because of the role of mitochondrial oxidative stress in CAD (Dikalov and Ungvari, 2013), it has been previously hypothesized that changes in mtDNAcn might be linked to hypertension (Prestes and Charchar, 2017). Nevertheless, only a few data on this topic have been collected until now. Lei et al. (Lei et al., 2017) measured mtDNAcn in white blood cells of a coal mining group located in northern China, and reported no differences between hypertensives and healthy controls. An elevated urinary mtDNAcn was detected in hypertensive patients, and it was correlated with markers of renal injury and dysfunction (Eirin et al., 2017, 2016). This increase might be explained considering that fragments of the mitochondrial genome released from dying cells are considered surrogate markers of mitochondrial injury. Moreover, mtDNAcn measured in urine samples was elevated in progressive acute kidney injury patients (Whitaker et al., 2015), and it was associated with glomerular hyperfiltration in obese African American hypertensive patients (Eirin et al., 2017). All these data support the involvement of mitochondrial dynamics in both the alteration of kidney function and hypertension. In our population, mtDNAcn decreased proportionally with GFR, in accordance with previous evidence suggesting that a higher blood mtDNA copy number is associated with a lower risk of incident chronic kidney disease (Tin et al., 2016). On the other hand, diabetes, that has been previously related to both the increase and decrease of mtDNAcn (Wong et al., 2009; Xu et al., 2012), was not significantly associated with mtDNAcn in our study.

In addition to classical risk factors for CVD, we also tested correlations between mtDNAcn and TMAO metabolism. While no associations with TMAO were measured, mtDNAcn directly correlated with TMA levels in the whole population. This finding suggests that a reduction of mitochondrial function might occur in individuals that display lower TMA. This is in accordance with our previously published data describing a subtle but significant reduction of TMA in CAD patients with respect to controls, while no significant differences were measured for TMAO between the two groups (Bordoni et al., 2020b).

A definite explanation of this phenomenon is still to be identified. In our cohort, a direct correlation between TMA and TMAO can be measured only in the CAD group, despite we did not observe a net increase of TMAO in CAD patients with respect to controls. Preliminary evidence suggested that TMA might have a role per se in CVD (Jaworska et al., 2019b, 2019a; Restini et al., 2020). Jaworska et al. showed that TMA was inversely correlated with the estimated glomerular filtration rate in humans; moreover, TMA reduced cardiomyocyte viability *in vitro*, while TMAO was even protective against TMA-induced cytotoxicity (Jaworska et al., 2019b). The authors suggested that the detrimental effect of TMA might be due to a triggering role of this molecule in the degradation of protein structures (i.e., albumin). The possibility of stable protein-TMA complexes (especially in the condition of increased oxidative stress, as occurring in inflammatory conditions (Bito et al., 2005; Era et al., 1995; Rahmani-Kukia et al., 2020)), together with the increased ratio of conversion of TMA into TMAO might provide a possible explanation for the low TMA levels measured in CAD patients and suggest a different fate and metabolism of TMA in CVD respect to healthy people. Studies further investigating this hypothesis are ongoing in our laboratories.

A limitation of this study is the absence of further data on urinary TMA and TMAO excretion. Moreover, although a positive correlation between the number of mitochondria and mtDNAcn has been demonstrated (Lee and Wei, 2000), mtDNAcn is not always a reliable predictor for mitochondrial abundance (Cayci et al., 2012; Lin et al., 2008; Liu et al., 2003; Qiu et al., 2013), probably because of the existence of compensatory effects. Thus, our study supports the involvement of TMA (but not TMAO) in CVD pathogenesis; nonetheless, further mechanistic investigations are necessary to clarify the real role of this metabolite in CVD and its usage as a clinical biomarker.

In this regard, we tested the possibility to predict CAD by using all measured variables, including mtDNAcn and other classical CVD risk factors. A logistic regression model identified decreased mtDNAcn, male gender, diagnosis of hypertension, smoke habits, and diagnosis of diabetes as significant risk factors for CAD development in this population; conversely, neither TMA nor TMAO have been included as significant predictors. The precision-recall analysis showed that it is possible to have a good prediction of CAD only by including information about mtDNAcn, gender and hypertension diagnosis (AUC = 0.894). Thus, validation in other cohorts of the usage of these 3 easy-to-measure parameters could promote their usage in population stratification for CVD risk.

Of note, cardiometabolic risk factors such as high blood pressure, obesity, and dyslipidemia have been previously shown to modify mtDNAcn (Hang et al., 2018; Huang et al., 2011). Thus, the hypothesis that environment and lifestyle factors might have major effects on mtDNAcn has been raised. Several pollutants (e.g., benzene (Carugno et al., 2012), particulate matter (Hou et al., 2013), and polycyclic aromatic hydrocarbons (Hou et al., 2010)) have been associated with changes in mtDNAcn. In addition, dietary factors such as fructose and salt can modify the mtDNAcn in blood (Hernández-Ríos et al., 2013). This is particularly interesting since we identified a correlation between mtDNAcn and hypertension. In conclusion, mtDNAcn appears as a plastic biomarker that might change in response to environmental and dietetic factors. Further research on modifiable CVD risk factors influencing mtDNA copy number may improve the prevention and treatment of this multifactorial pathology, making mtDNAcn a new tool to monitor complex disorders related to both metabolic imbalances and environmental insults at the molecular level (Torres, 2018).

## MATERIALS AND METHODS

### Study cohort recruitment and sample collection

CAD patients with angiographically confirmed CAD or with angina referred to elective or urgent coronary angiography have been recruited in Wejherowo Cardiovascular Center according to the following inclusion criteria. The diagnosis of myocardial infarction was established according to the Third Universal Definition of Myocardial Infarction, and angina was diagnosed according to the 2015 ESC guidelines for the management of NSTE-ACS3. CAD patients were treated with statins for a secondary prevention of cardiovascular morbidity. Control subjects without a self-reported medical history of CVD were recruited in the same region. The study was approved by the Regional Bioethical Committee (RBC) in Gdansk (KB-27/16 and KB 32-17) and registered at clinicaltrials.gov (NCT03899389). All methods were carried out in accordance with relevant guidelines and regulations approved by RBC. Informed consent was obtained from all subjects.

Venous blood samples were collected in EDTA-containing tubes. The plasma samples were prepared by centrifugation at 1300 g for 10 min at 18-25°C, and were kept frozen at −80°C.

### Assessment of variables related to cardiovascular risk

Plasma TMA and TMAO were determined by the Ultra-Performance Liquid Chromatography (UHPLC) tandem mass spectrometry method, as previously described (Bordoni et al., 2020b).

Body mass index (BMI) was calculated as weight in kilograms divided by height in meters squared. Hypertension and diabetes were diagnosed by medical doctors in diagnostic processes and were reported by subjects, as well as smoking habits (present or past), in self-reported questionnaires. GFR was calculated by Cockcroft-Gault equation (Cockcroft and Gault, 1976).

### MtDNAcn assessment

Genomic DNA was extracted from whole blood using the kit for genomic DNA purification (A&A Biotechnology, Gdynia, Poland). All samples have been processed under the same conditions and with the same DNA extraction method because variation of the DNA extraction method might affect the evaluation of mtDNAcn (Fazzini et al., 2018). Relative mtDNAcn quantification (Schmittgen and Livak, 2008) (considering nDNA as a normalizer), which is the current method of choice for mtDNAcn assessment (Malik and Czajka, 2013), was performed by real-time PCR (Biorad CFX96). Briefly, the cycle threshold (Ct) values of a mitochondrial-specific and nuclear-specific target were determined in triplicate for each sample. The difference in Ct values (ΔCt) for each sample represents a relative measure of mtDNcn. The following genes have been amplified for the detection of mitochondrial and nuclear DNA, respectively, using the listed primers: mtDNA-tRNALeu (fw: 5⍰-CACCCAAGAACAGGGTTTGT-3’; rv: 5⍰-TGGCCATGGGTATGTTGTTA-3’) for mitochondrial DNA, and beta-2-microglobulin (B2M) (fw: 5⍰-TGCTGTCTCCATGTTTGATGTATCT-3’; rv: 5⍰-TCTCTGCTCCCCACCTCTAAGT-3’) for the nuclear DNA. These primers have been verified by Fazzini and colleagues (Fazzini et al., 2018) for their specificity (unique amplification of mtDNA) and for the absence of co-amplified nuclear insertions of mitochondrial origin (NUMTs). An inter-run calibrator sample was used to adjust the results obtained from different amplification plates. All samples were anonymized for laboratory personnel.

### Statistical analysis

Power analysis for studying mtDNAcn in this population was performed according to the effect size reported by the meta-analysis from Yue and colleagues (Yue et al., 2018) and revealed a power>0.99. The Shapiro-Wilk test was used for the analysis of the normality of data distribution. Spearman correlation was calculated for testing the correlation among continuous variables. Bonferroni’s correction was applied to confirm statistical significance in multiple correlations. Chi-square test, Kruskal-Wallis test, and Generalized Linear Model (GLM) were used to test differences in the analyzed variables among groups adjusting for covariates. A stepwise logistic regression model was applied to identify (according to Wald statistics) which of the variables predicted the cardiovascular risk. Precision-recall curves were calculated to evaluate the performances of the prediction model as we are considering a database with unbalanced classes. Technical replicates are described in each specific material and methods section and are not considered for the inference statistics (no inflation of units of analysis was performed). Two-sided p values have been calculated, and significant differences were attributed to p values lower than 0.05.

## Data Availability

Data are available upon request.

## ABBREVIATION LIST

mtDNA: Mitochondrial DNA
mtDNAcn: Mitochondrial DNA copy number
CVD: Cardiovascular disease
CAD: Coronary artery disease
ATP: Adenosine triphosphate
BMI: Body Mass Index
TMA: Trimethylamine
TMAO: Trimethylamine-N-oxide
NUMT: Nuclear mitochondrial DNA sequence
nDNA: nuclear DNA

## AUTHOR CONTRIBUTIONS

Concept and design: LB, RG, RO; Study cohort recruitment: LL, JS, AS, RO; sample processing: IP, IP-M, AR; Statistical analysis: LB and MP; Drafting of the manuscript and data analysis: LB; Critical revision of the manuscript: RO, RG; Supervision: RG, LK, RO. All the authors approved the final version of the manuscript.

