## Supplementary materials for "Mitochondrial DNA copy number and trimethylamine levels in the blood: new insights on cardiovascular disease biomarkers"

Laura Bordoni

Unit of Molecular Biology, School of Pharmacy,

University of Camerino, Camerino,

62032

Italy

Tel. 0737/403211

^†^equal contribution

SUPPLEMENTARY MATERIALS

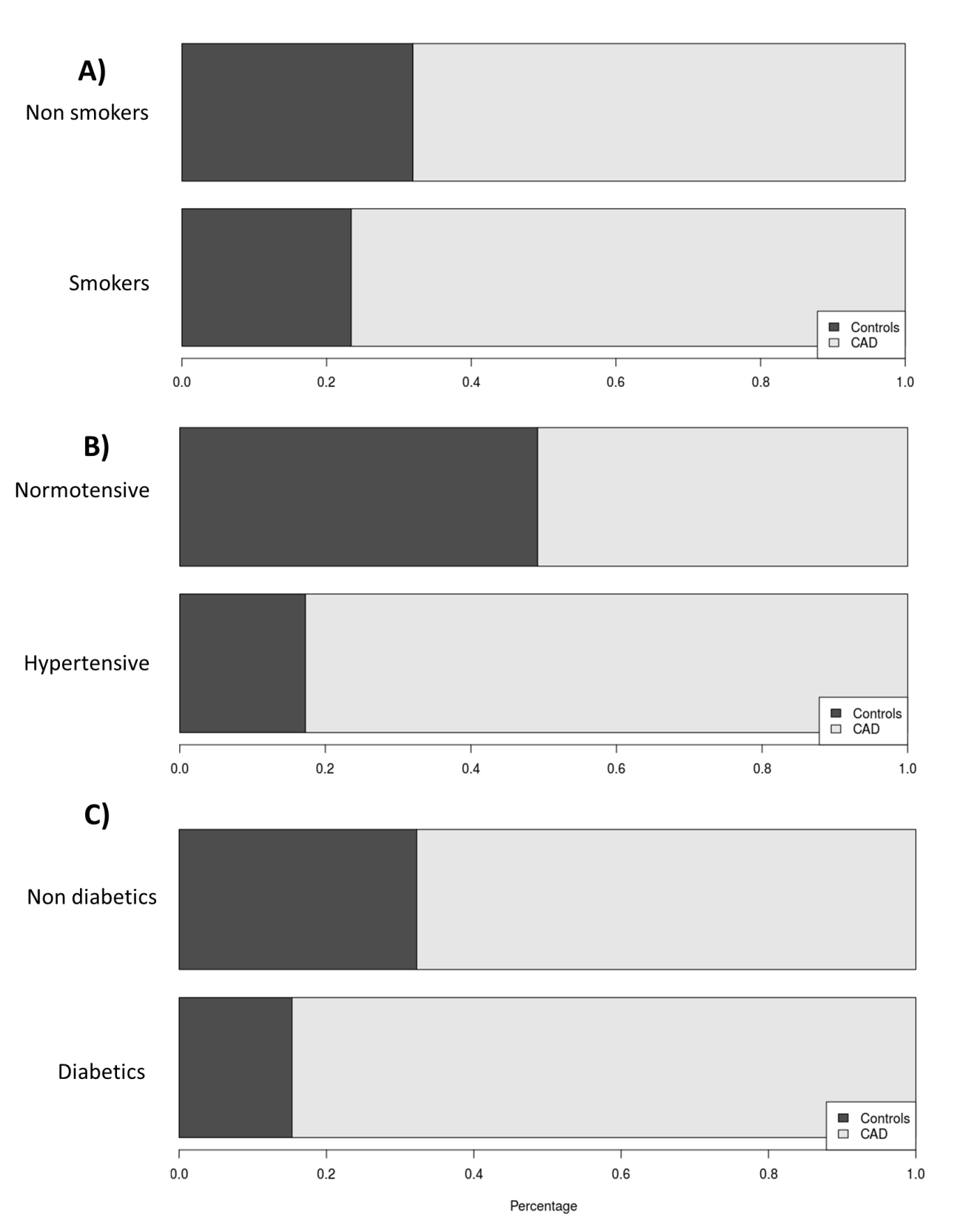

**Figure 1**. Smoking (A), hypertension (B) and diabetes (C) are confirmed as relevant risk factors for CAD in the analyzed population. A) pearson chi-quare. P=0.028; Odds Ratio [95%CI] = 1.535455 [ 1.047332 , 2.251073 ] B) pearson chi-quare. P=0.0001 Odds Ratio [95%CI] = 4.634116 [ 3.107077 , 6.911653 ] C) pearson chi-quare. P=0.0001 Odds Ratio [95%CI] = 2.630385 [ 1.580211 , 4.378483 ]

**Figure 2**

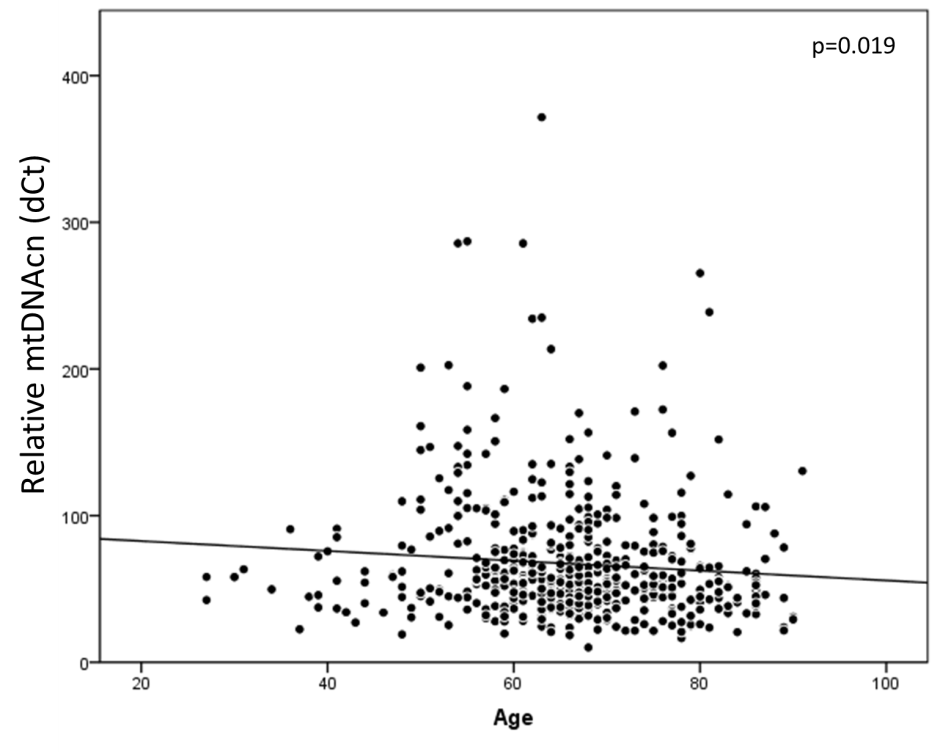
