## Supplementary figures and images for "Mitochondrial DNA copy number and trimethylamine levels in the blood: new insights on cardiovascular disease biomarkers"

### Supplemental figure 1

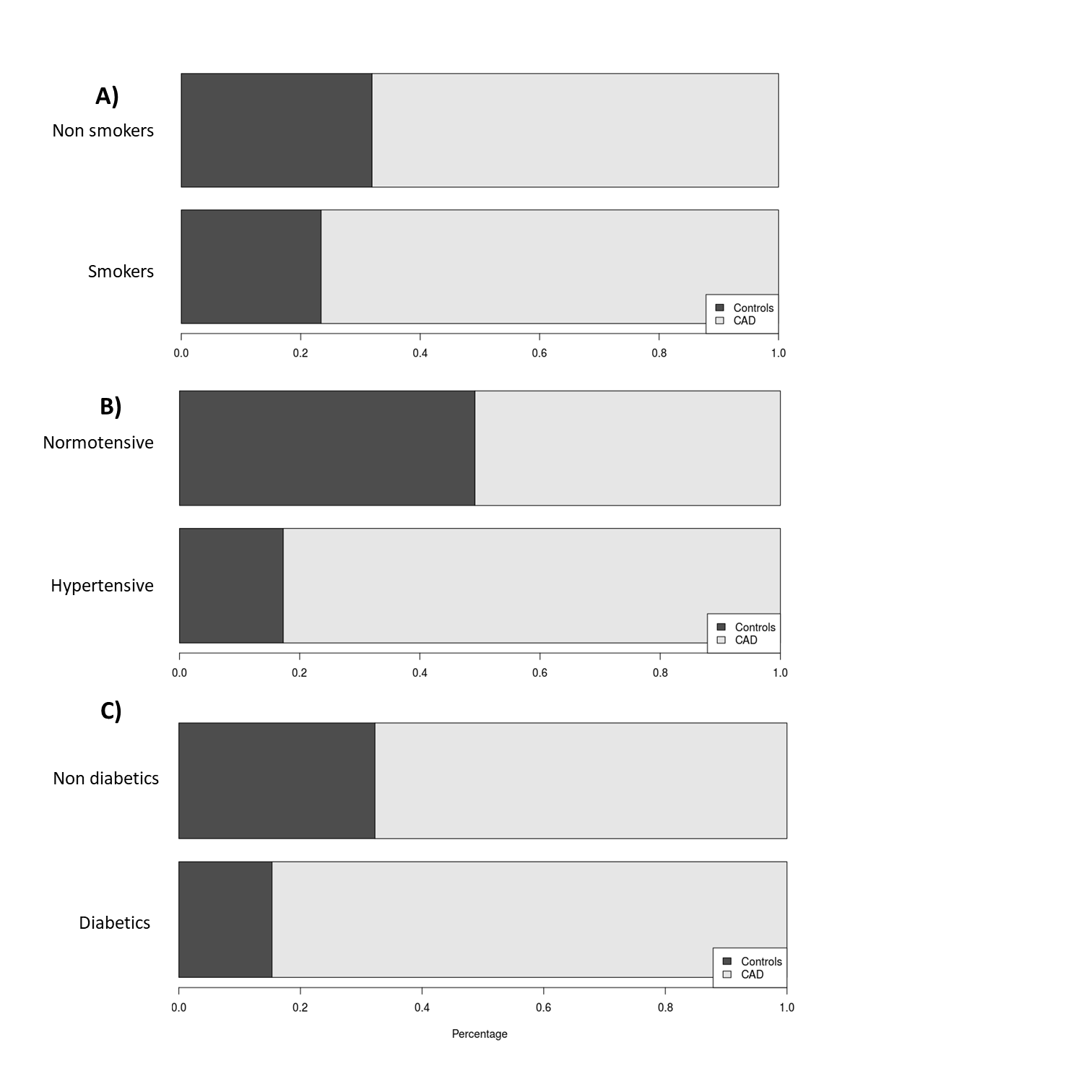

### Supplemental figure 2

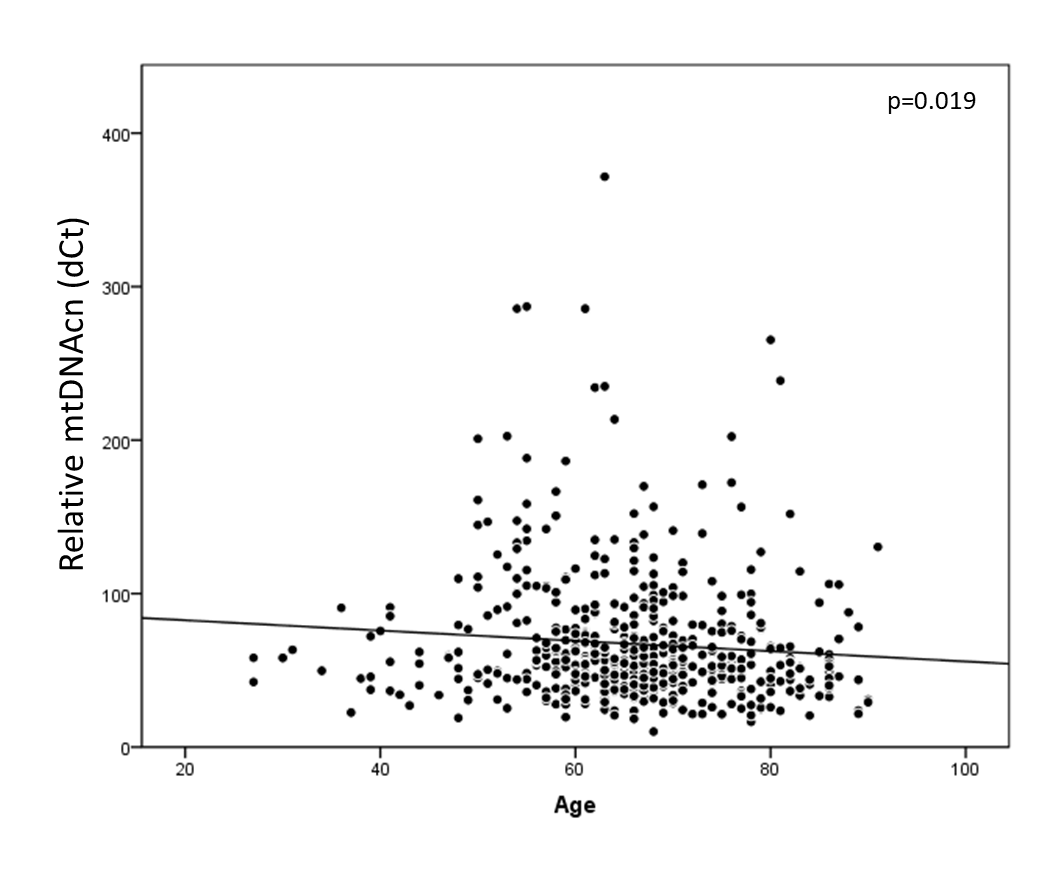
